# The role of the gluteofemoral adipose tissue in cerebrovascular disease risk: evidence from a mendelian randomization and mediation analysis

**DOI:** 10.1101/2024.08.08.24311685

**Authors:** Evangelos Pavlos Myserlis, Marios K. Georgakis, Livia Parodi, Ernst Mayerhofer, Jonathan Rosand, Chirantan Banerjee, Christopher D. Anderson

## Abstract

**Objective:** To explore causal associations between BMI-independent body fat distribution profiles and cerebrovascular disease risk, and to investigate potential mediators underlying these associations.

**Methods:** Leveraging data from genome wide association studies of BMI-independent gluteofemoral (GFAT), abdominal subcutaneous (ASAT), and visceral (VAT) adipose tissue volumes in UK Biobank, we selected variants associated with each trait, and performed univariable and multivariable mendelian randomization (MR) analyses on ischemic stroke and subtypes (large artery (LAS), cardioembolic (CES), small vessel (SVS)). We used coronary artery disease (CAD), carotid intima media thickness (cIMT), and an MRI-confirmed lacunar stroke as positive controls. For significant associations, we explored the mediatory role of four possible mediator categories in mediation MR analyses.

**Results:** Higher genetically proxied, BMI-independent GFAT volume was associated with decreased risk of ischemic stroke (FDR-p=0.0084), LAS (FDR-p=0.019), SVS (FDR-p<0.001), CAD (FDR-p<0.001), MRI-confirmed lacunar stroke (FDR-p=0.0053), and lower mean cIMT (FDR-p=0.0023), but not CES (FDR-p=0.749). Associations were largely consistent in pleiotropy- and sample structure-robust analyses. No association was observed between genetically proxied ASAT or VAT volumes and ischemic stroke/subtypes risk. In multivariable MR analyses, GFAT showed the most consistent independent association with ischemic stroke, LAS, and SVS. Common vascular risk factors were the predominant mediators in the GFAT-cerebrovascular disease axis, while adipose-tissue-specific adiponectin and leptin mediated a proportion of ischemic stroke and CAD risk.

**Interpretation:** Genetically proxied, BMI-independent higher GFAT volume is associated with reduced cerebrovascular disease risk. Although this is largely mediated by common vascular risk factor modification, targeting adipose-tissue specific pathways may provide additional cardiovascular benefit.

## Introduction

Adult obesity has an estimated prevalence of 42% in the U.S. and an estimated annual medical cost of nearly $173 billion U.S. dollars, with reports indicating increasing prevalence over the past decades^1-3^. Increased body weight has been associated with elevated cerebrovascular disease (CVD) risk^4^.

In order to effectively address obesity and its negative cerebrovascular outcomes, characterizing the phenotype becomes of crucial importance. While the body mass index (BMI) is a straightforward metric to quantify body composition and identify at-risk populations in clinical practice^3^, it can be limited in its ability to quantify CVD risk in certain populations who are classified as BMI-normal but have high-risk obesity characteristics such as abdominal fat deposition^5^. As such, alternative measures have been proposed, such as abdominal adiposity or the waist-to-hip ratio (WHR). Of note, observational data have indicated that WHR might be a better predictor of stroke risk compared to other adiposity metrics, including BMI^4^, an observation further supported by mendelian randomization (MR) analyses that have uncovered causal associations between genetically proxied WHR, but not BMI, with cerebrovascular disease risk^6^. Thus, it has become increasingly understood that body fat distribution, rather than total body fat, exerts a more prominent role in cerebrovascular disease risk prediction^6^. Furthermore, imaging-based assessments of body fat composition have shown that there is significant variability in the distribution of adipose tissue in the visceral versus the subcutaneous compartment within the same total body fat strata that differentially affects CVD risk^5^. Of note, genome-wide association studies (GWAS) have uncovered distinct genetic architectures and biological characteristics of the gluteofemoral (GFAT), abdominal subcutaneous (ASAT), and visceral adipose tissue (ASAT) distributions^7^, offering a more biologically-informed view of adiposity independent of BMI, and suggesting that divergent pathophysiologic processes may contribute to obesity’s detrimental role in cerebrovascular disease. Importantly, focusing on these different biological processes provides the opportunity for the discovery of novel adiposity-specific therapeutic interventions, as has been recently demonstrated by the emergence of combined glucose-dependent insulinotropic polypeptide (GIP) and glucagon-like peptide-1 receptor agonists (GLP-1 RAs) such as tirzepatide. These compounds, through their pleiotropic effects on anorexigenic, lipid-lowering, antihypertensive, and anti-inflammatory pathways beyond the anti-diabetic GLP1-RA-related effect, have emerged as promising agents for major cardiovascular event prevention^8^.

Mendelian randomization (MR) leverages the random reproductive allocation of genes in large populations to explore causal associations between modifiable risk factors and outcomes, limiting biases that may arise from observational data designs, such as reverse causation or confounding^9^. Apart from uncovering putative causal associations, MR has emerged as an important tool in dissecting intricate pathophysiological pathways that may contribute to various disease states^10^. In this study, we leverage large-scale genome-wide association data in combination with univariable and multivariable MR analyses to explore causal associations between biologically informed body fat distribution profiles, independent of BMI, and their association with cerebrovascular disease risk. We additionally explore the potential mediators that drive these associations, with the overarching aim to refine the link between obesity and stroke risk and to discover potential adiposity-related targets for intervention.

## Methods

### Study Design

A summary of our study design is provided in **Figure 1**. We used genetic variants from large-scale GWAS to proxy BMI-independent, MRI-derived GFAT, ASAT, and VAT distributions as exposure instruments in two-sample univariable MR designs on stroke and stroke subtypes and explored causal associations between these traits. We used three additional outcomes as positive controls: coronary artery disease (CAD), mean carotid intima media thickness (cIMT), and a more reliable, MRI-confirmed lacunar stroke phenotype from Traylor et al^11^. For significant associations between BMI-independent local fat distributions and the abovementioned outcomes, we explored the mediatory role of four different types of potential categories of mediators in two-step mediation MR analyses: common vascular risk factors (proxied by systolic blood pressure (SBP), type 2 diabetes mellitus (T2DM), and low-density lipoprotein (LDL)), insulin resistance (proxied by fasting insulin), whole-body inflammation (proxied by C-reactive protein (CRP)), and adipose-specific factors (proxied by adiponectin and leptin).

**Figure 1.**
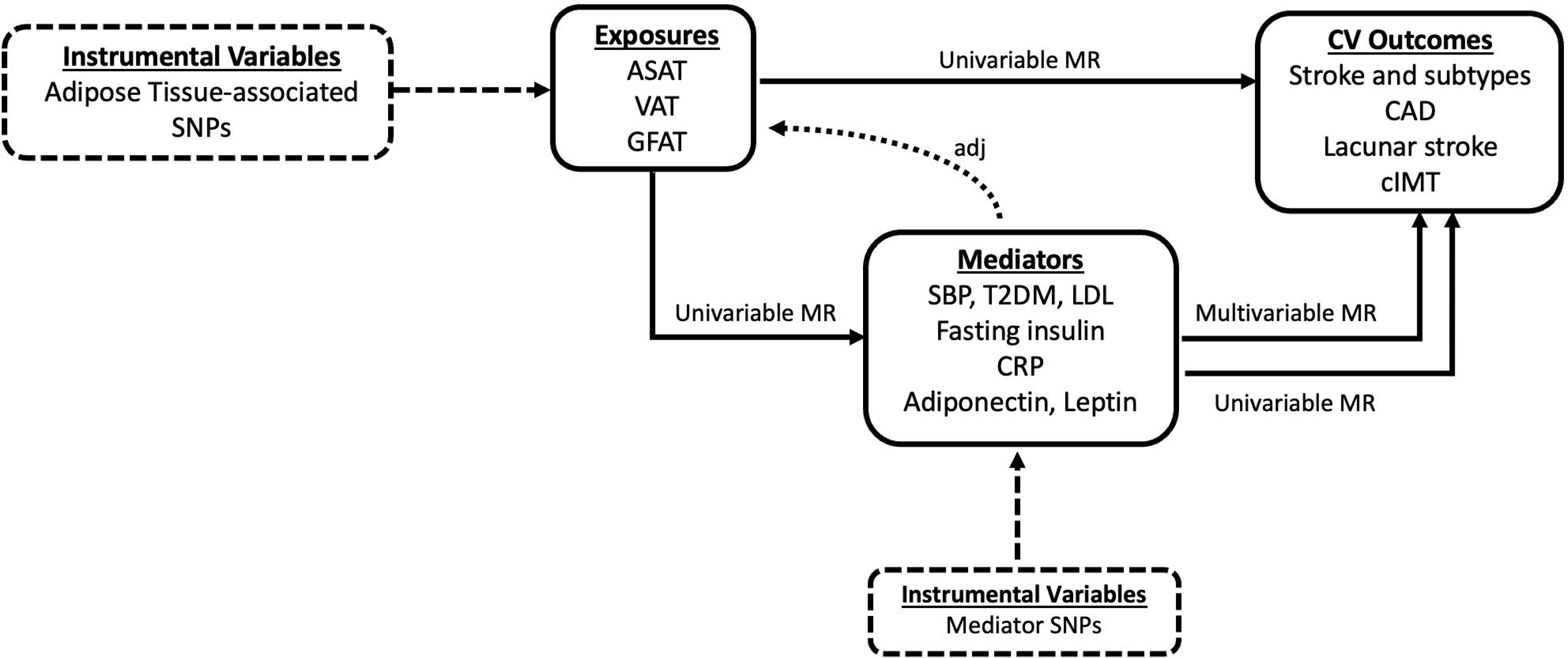
Study design. *SNPs single-nucleotide polymorphisms; ASAT abdominal subcutaneous adipose tissue; VAT visceral adipose tissue; GFAT gluteofermoral adipose tissue; CAD coronary artery disease; cIMT carotid intima media thickness; SBP systolic blood pressure; T2DM type 2 diabetes mellitus; LDL low-density lipoprotein; CRP c-reactive protein*

### Data sources and selection of genetic instruments

As genetic instruments, we selected variants from publicly available GWAS of relevant traits. We selected genome-wide (p < 5×10^-8^), independent (r^2^ < 0.01), BMI- and height-adjusted variants associated with GFAT, VAT, and ASAT from a GWAS of 38,965 UK Biobank participants with the above calculated MRI-derived adipose tissue volumes and available genotype array data, after quality control procedures^7^. GFAT, VAT, and ASAT volumes were derived using a convolutional neural network model from a subset of participants with available MRI data, and is described in detail elsewhere^12^. A brief description of the methodology used to generate the BMI-independent local adiposity profiles in UK Biobank and baseline characteristics is provided in the **Supplementary Methods**.

For ischemic stroke and stroke subtypes, we utilized GWAS data from the European participants of the GIGASTROKE consortium, which consisted of 62,100 cases of ischemic stroke (IS), 6,399 cases of large artery (LAS), 10,804 cardioembolic (CES), 6,811 small vessel stroke (SVS) and 1,234,808 controls^13^. We expanded our main analyses to three cardiovascular-related phenotypes that we selected as positive controls; CAD, MRI-confirmed lacunar stroke, and mean cIMT. For CAD, we utilized data from the CARDIoGRAMplusC4D consortium, comprising of 181,522 cases and 1,165,690 controls of predominately European (> 95%) ancestry^14^. For lacunar stroke, we leveraged data from the Traylor et al GWAS of MRI-confirmed lacunar strokes, that includes 6,030 cases and 248,929 controls of European ancestry^11^. In contrast to standard phenotyping that is based on the TOAST criteria, defined as a clinically-compatible lacunar stroke syndrome and the absence of other stroke causes or CT-based evidence of non-lacunar infarction, in this study, the authors gathered data from the UK DNA Lacunar Stroke studies 1 and 2, the International Stroke Genetics Consortium, as well as other prior studies, and centrally re-analyzed MRI images to confirm the presence of a lacunar infarction based on a set of prespecified criteria, thus providing a more detailed phenotyping of lacunes that helped uncover novel genomic loci not previously discovered. For mean carotid IMT, we utilized data from a UK Biobank GWAS study that included 45,185 participants^15^.

We explored four different categories of candidate mediators: 1) common vascular risk factors routinely addressed in everyday clinical practice (SBP, T2DM, and LDL), 2) insulin resistance (proxied by fasting insulin) given its independent association with cerebrovascular disease risk^16^, 3) CRP, given the recent anti-inflammatory agents’ emerging role in stroke risk prevention^17^, and 4) adipose-tissue specific factors (adiponectin, and leptin) to investigate whether obesity-specific factors might be contributing to stroke risk prediction. For SBP, we used GWAS data from a meta-analysis of UK Biobank and the International Consortium for Blood Pressure (ICBP) including 757,601 participants of European ancestry^18^. For T2DM, we used a GWAS of 80,154 cases and 853,816 controls of European ancestry in the DIAMANTE consortium^19^. For LDL, we used GWAS data from 1,320,016 European individuals in the Global Lipids Genetics Consortium (GLGC)^20^. For fasting insulin, we used GWAS data from ∼158,000 participants of European ancestry in the Meta-Analyses of Glucose and Insulin-related traits Consortium (MAGIC)^21^. For CRP, we leveraged data from European participants of a CRP GWAS meta-analysis of UK Biobank and the Cohorts for Heart and Aging Research in Genomic Epidemiology (CHARGE) Consortium^22^. For adiponectin, we utilized GWAS data from 2,962 men in the Metabolic Syndrome in Men (METSIM) study^23^, and for leptin, we used data from an exome-based analysis of leptin concentrations in 49,909 individuals of European ancestry^24^. A summary of our data sources is depicted in **Table 1**.

**Table 1.**
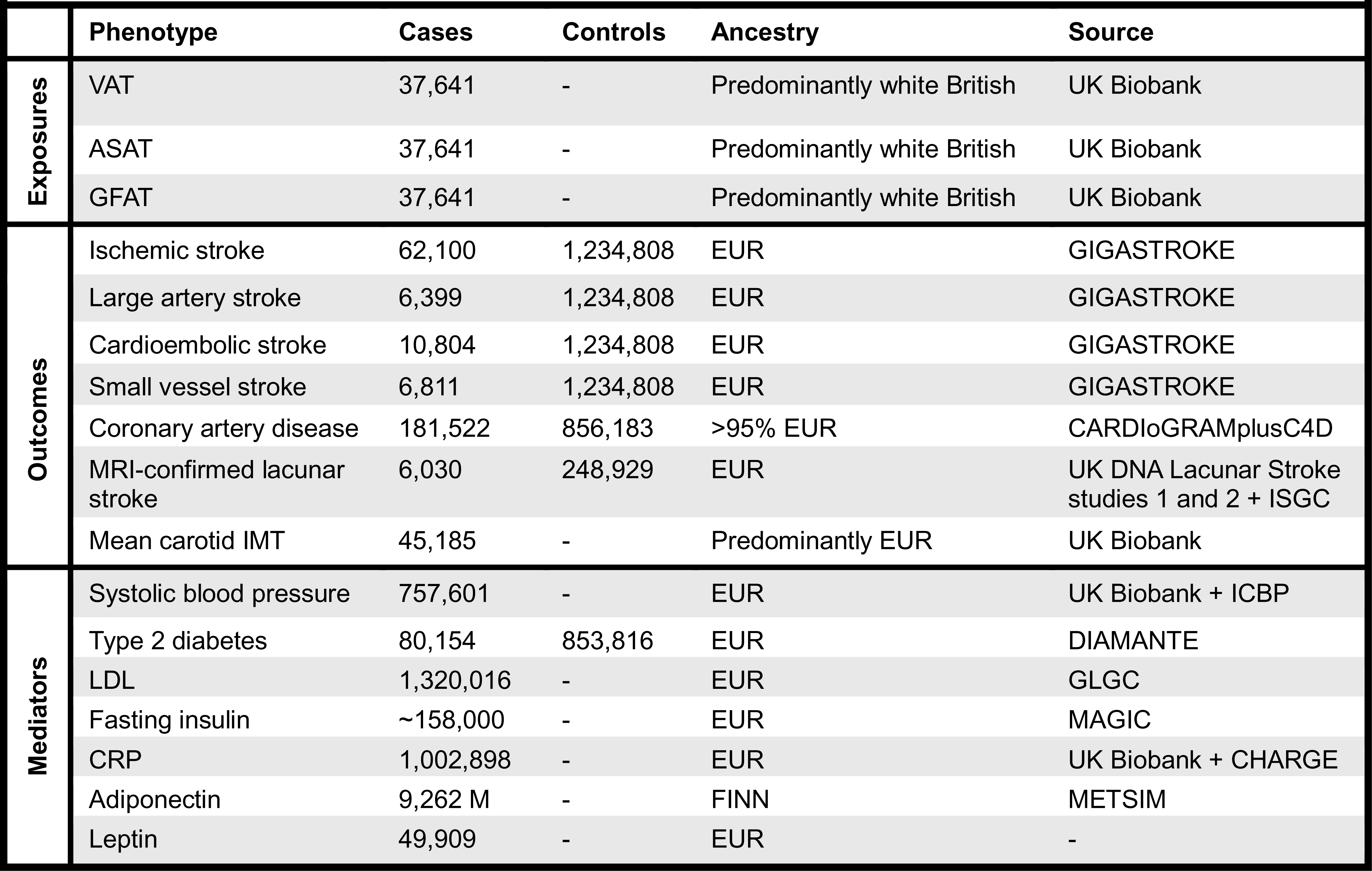
Characteristics of included studies. *VAT visceral adipose tissue; ASAT abdominal subcutaneous adipose tissue; GFAT gluteofemoral adipose tissue; EUR European; FINN Finnish; CARDIoGRAMplusC4D coronary artery disease genome wide replication and meta-analysis (CARDIoGRAM) plus the coronary artery disease (C4D) genetics; ISGC international stroke genetics consortium; IMT intima media thickness; DIAMANTE diabetes meta-analysis of trans-ethnic association studies; LDL low density lipoprotein; CRP c-reactive protein; ICBP international consortium for blood pressure; GLGC global lipids genetics consortium; MAGIC meta-analyses of glucose and insulin-related traits consortium; cohorts for heart and aging research in genomic epidemiology; METSIM metabolic syndrome in men; M male*

### Statistical Analyses

#### Main mendelian randomization analyses

We performed two-sample, univariable inverse-variance weighted (IVW) MR with the main outcomes, ischemic stroke and stroke subtypes (LAS, CES, and SVS)^25^. Heterogeneity was evaluated using the *I^2^* index, Cochran’s *Q* statistic, and Cochran’s *Q* p-value (significant if < 0.05) ^26^. For significant IVW associations, we conducted sensitivity analyses with alternative MR approaches to account for pleiotropic effects. We specifically utilized the weighted median estimator, which requires at least half of the variants in the genetic instruments to be valid^27^, and Egger regression, which allows up to all genetic variants to have pleiotropic effects, as long as they are not proportional to the variants’ effects on the exposure of interest^28^. We also used the MR-Egger intercept to evaluate for evidence of horizontal pleiotropy (significant if p < 0.05).

Given the potential sample overlap that may exist between large datasets of the included phenotypes, raising the possibility of inflated type 1 errors and biased MR estimates, we additionally utilized an MR technique, MR-APSS, that accounts for sample structure between exposure-outcome datasets^29^. MR-APSS uses a proposed background-foreground model to evaluate for causal effects; the background model accounts for correlated pleiotropy and sample structure (including population stratification, cryptic relatedness, and sample overlap) utilizing genome-wide GWAS data, while the foreground model evaluates for a causal effect under the InSIDE (Instrument Strength Independent of Direct Effect) assumption, after correlated pleiotropy has been accounted for in the background model. Furthermore, this method is insensitive to p-value threshold selection for the generation of the genetic proxy, providing non-inflated type 1 error rates with addition of more invalid single nucleotide polymorphisms (SNPs), while statistical power is improved with the use of more lenient thresholds. A p value threshold of 5×10^-5^ is recommended in real-world settings^29^. In our analyses, we utilized p=5×10^-5^, kb=10000, and r^2^<0.01 as parameters for instrument selection, however we performed an additional analysis with a p-value threshold of 5×10^-8^ (while keeping the clumping window and r^2^ the same) to maintain consistency across MR methodologies. Sample sizes used for the MR-APSS analyses are those depicted in **Table 1** (sample size or number of cases/controls, for continuous or case/control traits, respectively).

When a significant univariable MR association was found between an exposure and a main outcome, we performed two additional analyses: reverse MR to explore the possibility of reverse causation between an exposure and a main outcome, and multivariable MR to investigate the independent effect of each BMI-independent local adiposity phenotype on that outcome. For reverse MR, we first generated genetic instruments for ischemic stroke and stroke subtypes by extracting genome-wide significant (p < 5×10^-8^), independent (r^2^ < 0.01) variants from the aforementioned GWAS to proxy these traits. We then performed univariable IVW MR on the respective exposures. For multivariable MR, we followed the following analytical steps: first, we combined all SNPs from the three genetic instruments for GFAT, ASAT, and VAT, and extracted the unique SNPs. Then, we re-clumped the variants using an r^2^ threshold of < 0.01 to derive independent SNPs, and harmonized all effect size estimates according to GFAT. Lastly, we ran multivariable MR on our main outcomes utilizing the multivariable IVW^30^, median-based, and MR-Egger^31^ approaches.

For univariable MRs, all SNPs and effect estimates were harmonized across datasets to the effect allele of the exposure datasets, or to the effect allele of the outcome datasets (ischemic stroke or subtypes) in the case of reverse MR. The F-statistic was used to evaluate instrument strength^32^. If the F-statistic was higher than 10, indicating strong instrument selection, then weak instrument bias that might have been introduced by violating the two-sample assumption stemming from potentially using overlapping samples between exposures and outcomes was less likely^33^. For each exposure instrument, we also calculated the phenotypic variance explained (R^2^) using the methodology described in the **Supplementary Methods**^34^. Given that in the primary GWAS analysis of GFAT, ASAT, and VAT, each adiposity trait was inverse-normal transformed before conducting GWAS^7^, MR estimates were expressed as per standard deviation (SD) increase in adjusted adipose tissue volume. For significant univariable associations, we also performed main IVW and sensitivity MR analyses on our positive controls, CAD, MRI-confirmed lacunar stroke, and mean carotid IMT. The false discovery rage (FDR) was used to correct for multiple hypothesis testing across the main outcomes and positive controls for each univariable MR method separately^35^.

#### Mediation analyses

For significant associations in the main analyses, we conducted two-step mediation MR analyses to explore possible mediators^10^. We considered four different categories: common vascular risk factors (SBP, T2DM, LDL), insulin resistance (fasting insulin), systemic inflammation (CRP), and adipose-tissue specific factors (circulating adiponectin and leptin). We first performed two-sample univariable IVW MR between the exposure and the mediators. Then, we performed univariable MR between mediators and outcomes, after generating genetic instruments for our mediators, similar to the above-described methodology, i.e. after extracting genome-wide (p < 5×10^-8^), independent (r^2^ < 0.01 for all mediators apart from adiponectin (r^2^ < 0.1) in the setting of low sample size), variants associated with each mediator. For significant univariable MR associations, we performed multivariable MR of the mediator on the outcome, after adjusting for the effect of the mediator genetic instrument on the exposure. We calculated the indirect effect of the mediator by multiplying the univariable MR estimate between the exposure and the mediator with the multivariable MR estimate between the mediator and the outcome. Lastly, we calculated the percent mediated after dividing the indirect effect by the overall effect of the exposure on the outcome. Confidence intervals for the percent mediated were calculated after bootstrapping over 1000 iterations. A schematic of our workflow is depicted in **Figure 1**. We considered three different types of mediators: candidate mediators were those that showed significant univariable MR associations with both the exposure and the outcome; suggestive and significant mediators were those with p value between 0.1 and 0.05, and less than 0.05 in multivariable MR, respectively. All analyses were performed in R studio version 3.6.1, using the MendelianRandomization^36^, TwoSampleMR^37, 38^, and MR-APSS^29^ packages^39^.

## Results

### Main outcomes

We extracted 44 SNPs for BMI- and height-adjusted GFAT, 19 for ASAT, and 25 for VAT volumes, respectively. All SNPs had an F value > 10, indicating strong genetic instrument selection. The phenotypic variance explained (R^2^) by each instrument was ∼5.98%, ∼2.07%, and ∼3.3% for GFAT, ASAT, and VAT, respectively **(Supplementary Tables 1-3)**. In univariable MR analyses, one SD increase in genetically proxied BMI-independent GFAT volume was associated with decreased risk of ischemic stroke (OR 0.92; 95% confidence interval (CI) 0.86-0.98; FDR-p=0.0084). Higher genetically predicted GFAT volume was also associated with decreased risk of LAS and SVS (OR 0.80 per SD increase, 95% CI 0.66-0.96, FDR-p=0.019; and OR 0.77 per SD increase, 95% CI 0.67-0.88, FDR-p<0.001, respectively), but not CES (OR 0.98 per SD increase, 95% CI 0.87-1.11, FDR-p=0.749) (**Fig 2A**). There was evidence of significant heterogeneity in the MR estimates for ischemic stroke and LAS (*I^2^*=45.5%, *Q*=67.8768, *Q* p-value=0.0015; and *I^2^*=38.6%, *Q*=53.7332, *Q* p-value=0.0127, respectively), but not for SVS (*I^2^*=0.0%, *Q*=28.7836, *Q* p-value=0.6772), whereas there was no evidence of horizontal pleiotropy in all estimates (MR Egger intercept p-values > 0.05). In weighted median analyses, effect size estimates remained directionally consistent and largely significant, whereas in MR Egger analyses, effect sizes were even larger, although not significant, probably due to the lower statistical power of this method (**Supplementary Table 4**). In MR-APSS analyses, effect size estimates remained directionally concordant and statistically significant, across ischemic stroke and stroke subtypes **(Supplementary Table 4)**. Apart from a suggestive reverse MR association between LAS and GFAT which was characterized by significant heterogeneity (Cochran’s Q p-value = 0.019), there was no other evidence that genetic predisposition to stroke or stroke subtypes was causally associated with GFAT in reverse MR analyses (no instrument could be built for SVS given that there were no genome-wide significant variants) (**Supplementary Table 5**). No association was observed between genetically proxied BMI- and height-adjusted ASAT or VAT volumes and ischemic stroke or stroke subtypes risk (**Fig 2A, Supplementary Table 6**). When all three local adiposity profiles were included in a multivariable MR model, GFAT volume showed the most consistent independent association with ischemic stroke, LAS, and SVS (**Fig 2B, Supplementary Table 7**). VAT also showed an independent association with ischemic stroke in multivariable IVW- and MR-Egger-based analyses, although the results suffered from significant heterogeneity (*Q* p-value 0.0001 and < 0.001, respectively).

**Figure 2.**
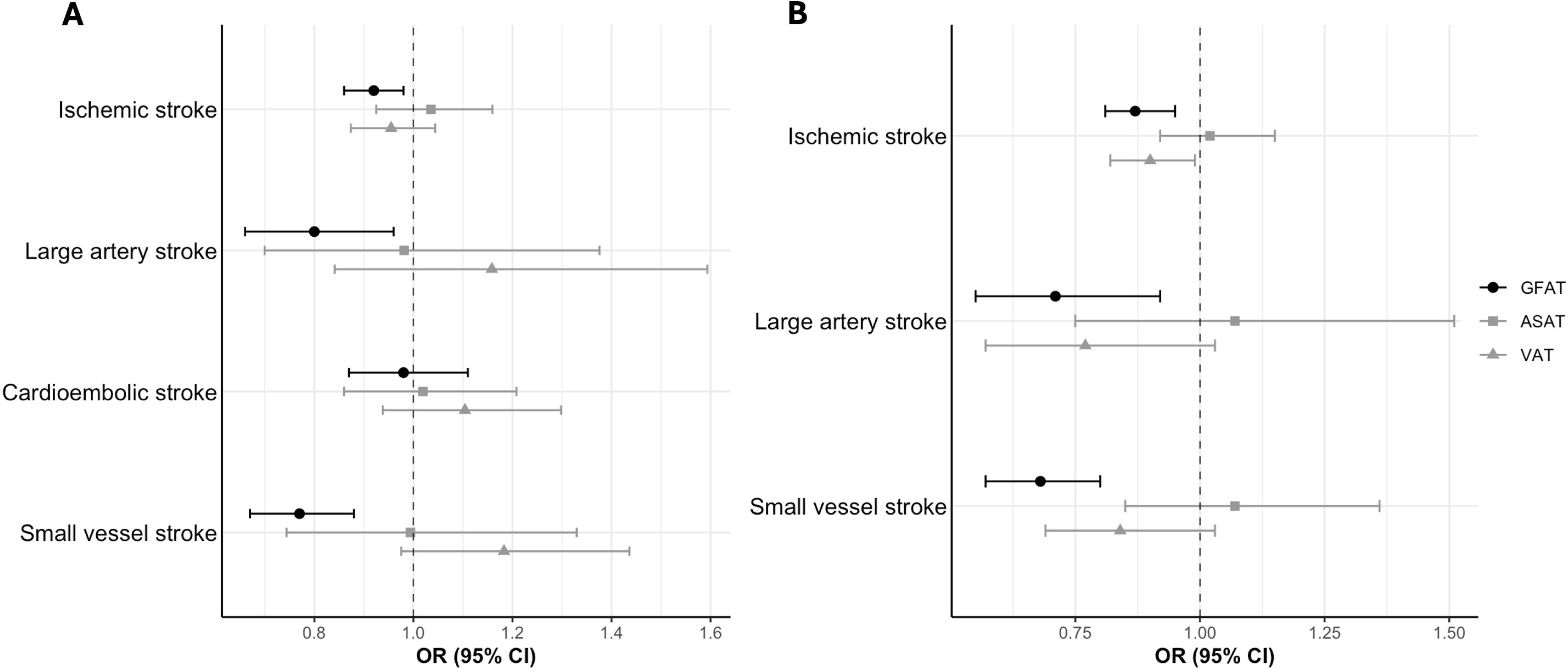
(A) Univariable and (B) multivariable inverse-variance weighted MR associations between BMI-adjusted local adiposity profiles and ischemic stroke and stroke subtypes.

### Relevant vascular phenotypes

In the setting of the observed protective association between BMI-independent GFAT volume and ischemic stroke and stroke subtypes risk, we performed follow-up analyses on relevant vascular phenotypes, CAD, MRI-confirmed lacunar stroke, and cIMT. We found that genetically proxied GFAT volume was associated with a decreased risk for CAD and MRI-confirmed lacunar stroke (OR 0.82 per SD increase, 95% CI 0.76-0.88, FDR-p<0.001, and OR 0.78 per SD increase, 95% CI 0.67-0.92, FDR-p=0.0053, respectively), as well as lower mean cIMT (beta=- 0.073 per SD increase, 95% CI (-0.114) - (-0.031), FDR-p=0.0023). Associations remained directionally concordant in sensitivity analyses, while a non-significant effect was observed between genetically predicted GFAT volume and mean cIMT in MR-APSS analysis with the more stringent 5×10^-8^ p-value threshold for instrument selection, possibly owning to the lower statistical power of this approach (**Fig 3, Supplementary Table 8**).

**Figure 3.**
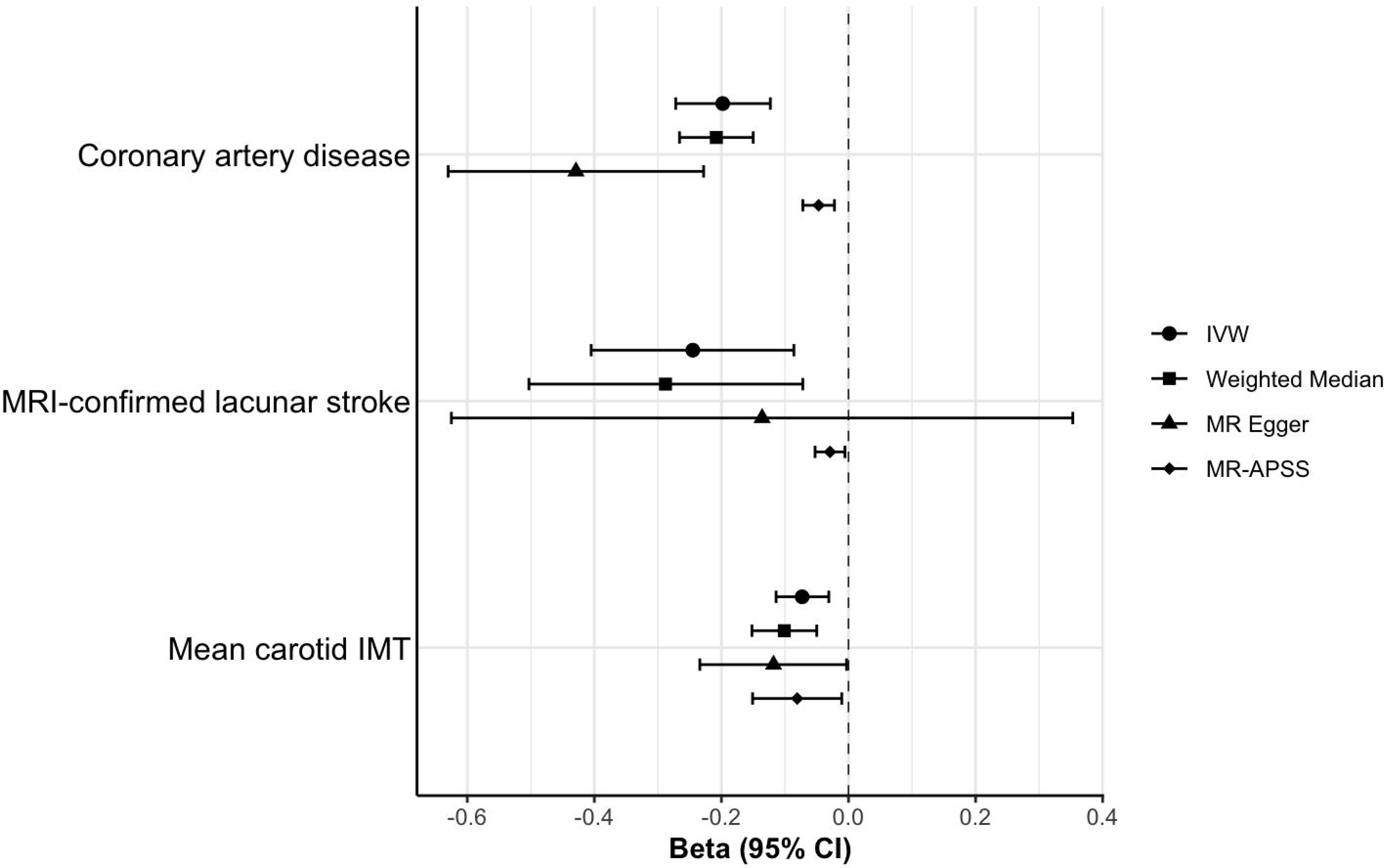
Association between BMI-independent GFAT volume and relevant vascular phenotypes. *IMT carotid intima media thickness; IVW inverse-variance weighted; CI confidence interval; GFAT gluteofemoral adipose tissue*

### Mediation analyses

We explored the potential mediation effect of the four categories of mediators between BMI- and height adjusted GFAT and the main outcomes and relevant vascular phenotypes. **Table 2** summarizes the results of the mediation analyses. Overall, the common vascular risk factors were the predominant mediators across phenotypes. Specifically, GFAT exerted a protective effect on all outcomes through lower systolic blood pressure. It also had a protective role on all outcomes except mean carotid IMT through reduction in type 2 diabetes risk, and resulted in lower risk for all outcomes except small vessel stroke and MRI-confirmed lacunar stroke through lower LDL levels. Neither insulin resistance, nor systemic inflammation, as proxied by fasting insulin and CRP respectively, were mediators in the GFAT-cardiovascular risk axis. With regards to adipose-tissue specific factors, adiponectin emerged as a significant mediator in ischemic stroke and suggestive mediator in CAD, two of the most well-powered outcomes. Additionally, leptin was a candidate mediator in CAD (**Table 2**, **Supplementary Tables 9-11**) While common vascular risk factors mediated the majority of the proportion across phenotypes (9-58% for SBP, 6-40% for T2DM, 3-13% for LDL), adiponectin mediated 14.9% (1.8% - 57%) of ischemic stroke and 4.6% (0.8% - 13.5%) of CAD risk, respectively (**Fig 4, Supplementary Table 12**). Details of genetic instruments for the mediators are reported in **Supplementary Tables 13-18**.

**Figure 4.**
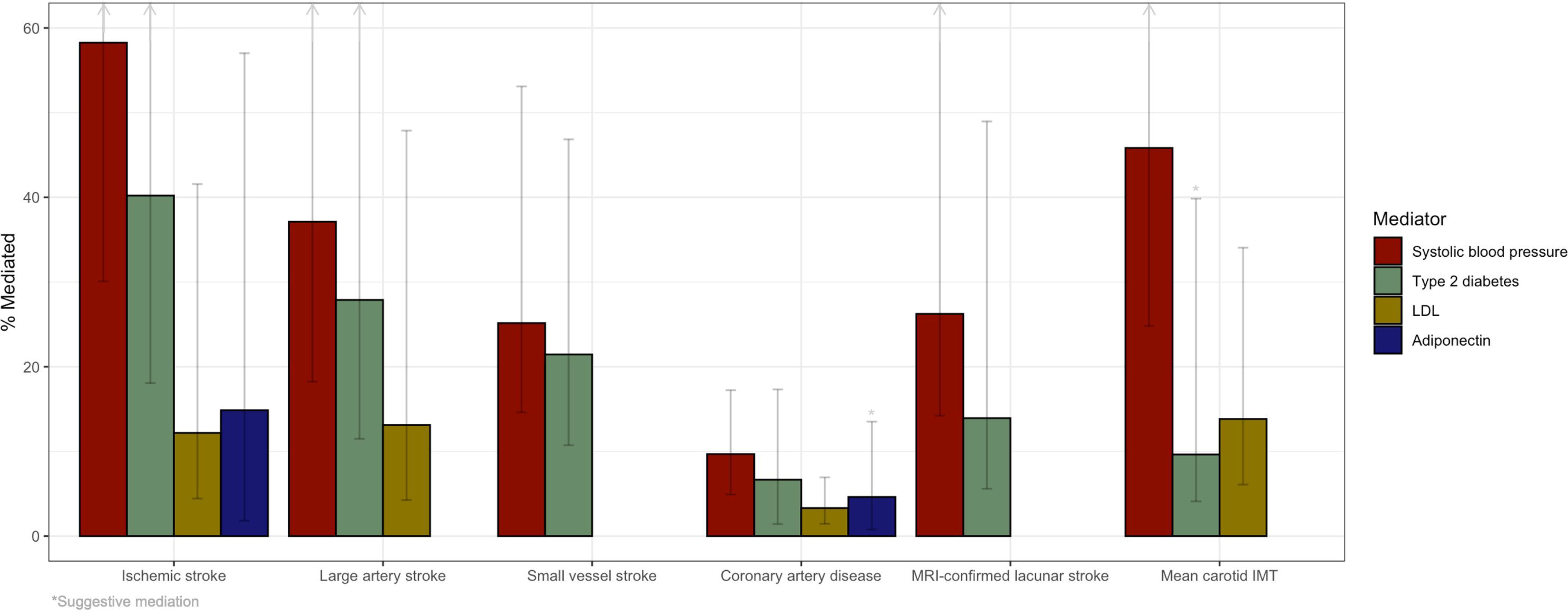
Percentage mediated by suggestive or significant mediators across outcomes. *IMT intima media thickness*

**Table 2.**
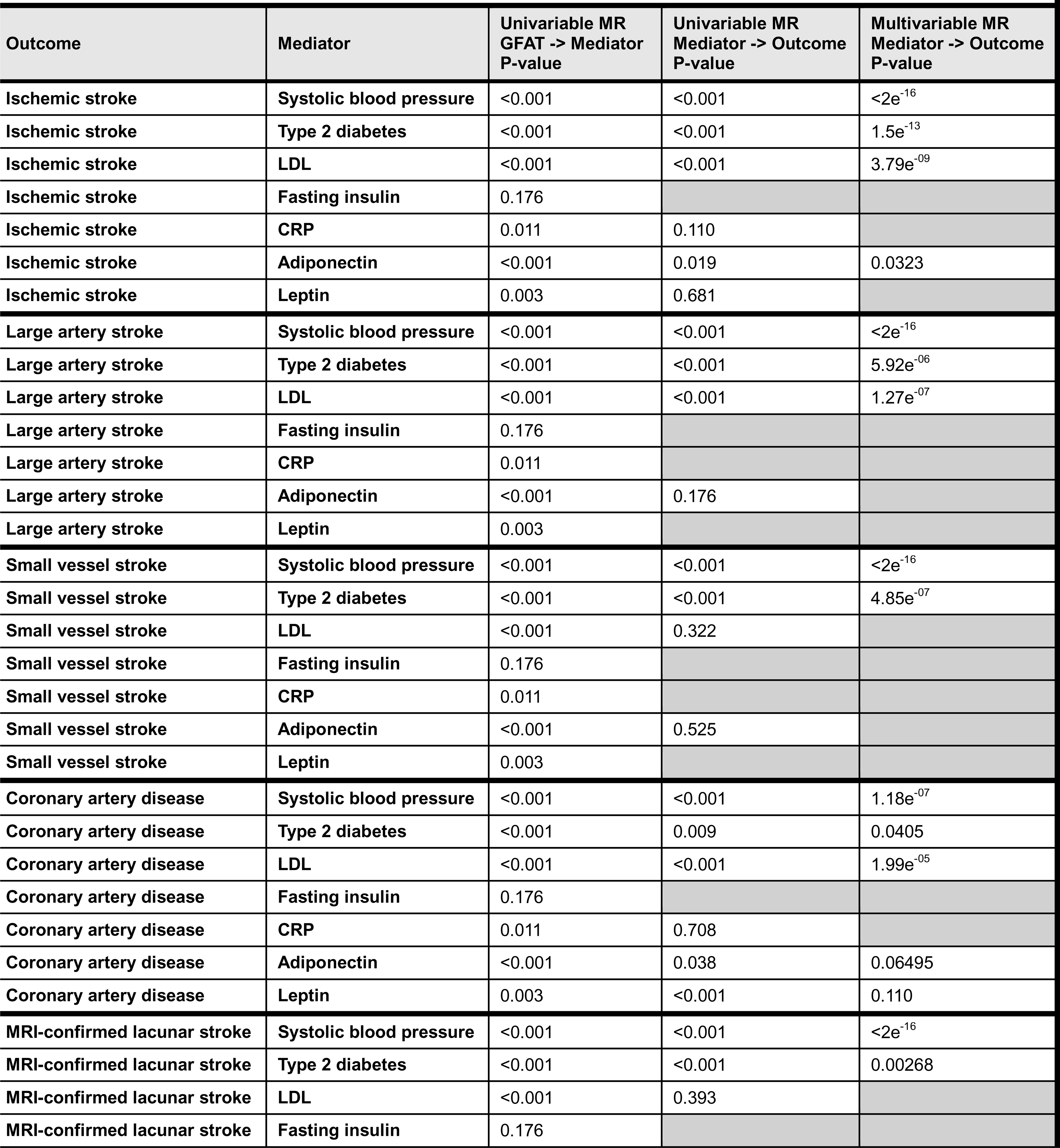
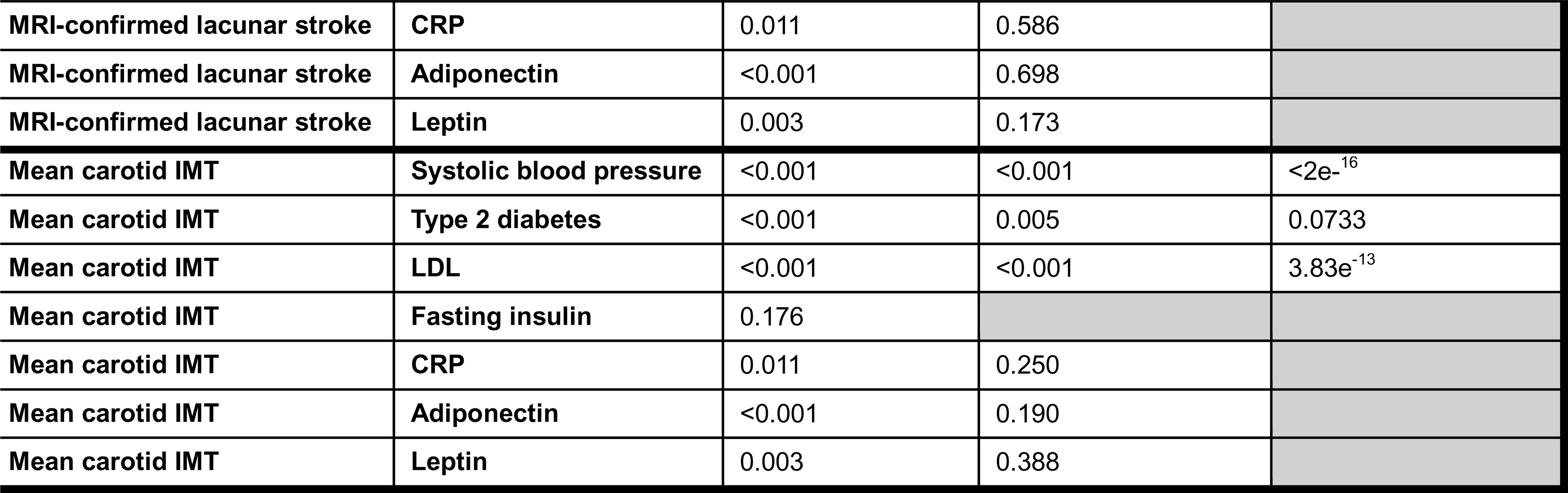
Results of mediation analysis. *MR mendelian randomization; GFAT gluteofemoral adipose tissue; LDL low density lipoprotein; CRP c-reactive protein; IMT intima media thickness*

| Outcome | Mediator | Univariable MR<br>GFAT -> Mediator<br>P-value | Univariable MR<br>Mediator -> Outcome<br>P-value | Multivariable MR<br>Mediator -> Outcome<br>P-value |
| --- | --- | --- | --- | --- |
| Ischemic stroke | Systolic blood pressure | <0.001 | <0.001 | <2e<sup>-16</sup> |
| Ischemic stroke | Type 2 diabetes | <0.001 | <0.001 | 1.5e<sup>-13</sup> |
| Ischemic stroke | LDL | <0.001 | <0.001 | 3.79e<sup>-09</sup> |
| Ischemic stroke | Fasting insulin | 0.176 |  |  |
| Ischemic stroke | CRP | 0.011 | 0.110 |  |
| Ischemic stroke | Adiponectin | <0.001 | 0.019 | 0.0323 |
| Ischemic stroke | Leptin | 0.003 | 0.681 |  |
| Large artery stroke | Systolic blood pressure | <0.001 | <0.001 | <2e<sup>-16</sup> |
| Large artery stroke | Type 2 diabetes | <0.001 | <0.001 | 5.92e<sup>-06</sup> |
| Large artery stroke | LDL | <0.001 | <0.001 | 1.27e<sup>-07</sup> |
| Large artery stroke | Fasting insulin | 0.176 |  |  |
| Large artery stroke | CRP | 0.011 |  |  |
| Large artery stroke | Adiponectin | <0.001 | 0.176 |  |
| Large artery stroke | Leptin | 0.003 |  |  |
| Small vessel stroke | Systolic blood pressure | <0.001 | <0.001 | <2e<sup>-16</sup> |
| Small vessel stroke | Type 2 diabetes | <0.001 | <0.001 | 4.85e<sup>-07</sup> |
| Small vessel stroke | LDL | <0.001 | 0.322 |  |
| Small vessel stroke | Fasting insulin | 0.176 |  |  |
| Small vessel stroke | CRP | 0.011 |  |  |
| Small vessel stroke | Adiponectin | <0.001 | 0.525 |  |
| Small vessel stroke | Leptin | 0.003 |  |  |
| Coronary artery disease | Systolic blood pressure | <0.001 | <0.001 | 1.18e<sup>-07</sup> |
| Coronary artery disease | Type 2 diabetes | <0.001 | 0.009 | 0.0405 |
| Coronary artery disease | LDL | <0.001 | <0.001 | 1.99e<sup>-05</sup> |
| Coronary artery disease | Fasting insulin | 0.176 |  |  |
| Coronary artery disease | CRP | 0.011 | 0.708 |  |
| Coronary artery disease | Adiponectin | <0.001 | 0.038 | 0.06495 |
| Coronary artery disease | Leptin | 0.003 | <0.001 | 0.110 |
| MRI-confirmed lacunar stroke | Systolic blood pressure | <0.001 | <0.001 | <2e<sup>-16</sup> |
| MRI-confirmed lacunar stroke | Type 2 diabetes | <0.001 | <0.001 | 0.00268 |
| MRI-confirmed lacunar stroke | LDL | <0.001 | 0.393 |  |
| MRI-confirmed lacunar stroke | Fasting insulin | 0.176 |  |  |
| **MRI-confirmed lacunar stroke** | **CRP** | 0.011 | 0.586 |  |
| **MRI-confirmed lacunar stroke** | **Adiponectin** | <0.001 | 0.698 |  |
| **MRI-confirmed lacunar stroke** | **Leptin** | 0.003 | 0.173 |  |
| **Mean carotid IMT** | **Systolic blood pressure** | <0.001 | <0.001 | <2e<sup>-16</sup> |
| **Mean carotid IMT** | **Type 2 diabetes** | <0.001 | 0.005 | 0.0733 |
| **Mean carotid IMT** | **LDL** | <0.001 | <0.001 | 3.83e<sup>-13</sup> |
| **Mean carotid IMT** | **Fasting insulin** | 0.176 |  |  |
| **Mean carotid IMT** | **CRP** | 0.011 | 0.250 |  |
| **Mean carotid IMT** | **Adiponectin** | <0.001 | 0.190 |  |
| **Mean carotid IMT** | **Leptin** | 0.003 | 0.388 |  |

## Discussion

In this study we leveraged large-scale genomic data to explore causal associations between biologically informed BMI-independent local adiposity distribution profiles and cerebrovascular disease risk. We found that, preferentially distributed adipose tissue in the gluteofemoral area, independent of BMI, is causally associated with decreased ischemic stroke and relevant vascular phenotype risk. No causal associations were found for BMI-adjusted VAT or ASAT. We also found that common vascular risk factors, typically addressed in everyday clinical practice, are predominant mediators of the association between GFAT and our outcomes. However, residual protective effect on vascular risk may be mediated by adipose-tissue specific factors, such as common adipokines adiponectin and leptin. Our results provide several important insights related to the pathophysiology of obesity-related cardiovascular disease risk that could inform future therapeutic strategies.

First, we provide evidence that GFAT distribution independent of BMI is causally associated with reduced risk of ischemic stroke and cardiovascular disease risk. This is in line with previous studies that have uncovered the favorable role of GFAT in type 2 diabetes liability and CAD risk^12^, and lends further support to the hypothesis of a GFAT-driven cardiometabolically healthy body fat distribution that is independent of overall weight or BMI^40^. Conversely, we found no association of BMI-adjusted VAT or ASAT with ischemic stroke in univariable MR analyses. Overall, these findings support the hypothesis that increases in stroke risk in the setting of an unhealthy body fat distribution (that is, adipose tissue not preferentially distributed in the gluteofemoral area) may primarily be driven by an inability of the gluteofemoral fat to expand, rather than relative increases in ASAT or VAT volumes^7^. The hypothesis that subcutaneous fat may act as an energy “buffer” in the setting of either excessive energy intake or insufficient spending is supported by prior animal as well as human studies. For instance, in a study by Gavrilova et al., the subcutaneous transplantation of wild-type fat in transgenic A-ZIP/F-1 lipoatrophic mice led to improvement in several metabolic markers, such as glucose levels, insulin sensitivity, and decreased weight as well as decreased hepatic size, steatosis, and triglyceride levels^41^. Similarly, data from human studies have suggested that antidiabetic treatment with thiazolidinediones – activators of PPARγ receptors most commonly found in subcutaneous rather than visceral fat – is associated with an increase in subcutaneous adipose tissue and a decrease in visceral fat, correlating with improvement in glycemic markers such as HbA1c and fasting plasma glucose^42^. Taken together, these studies support the hypothesis that preferential GFAT deposition over VAT or ASAT, independent of overall BMI, may lie in a causal protective cardiometabolic state, as suggested by our results.

Second, we found that the protective role of GFAT distribution in cardiovascular disease risk is primarily mediated by common vascular risk factors, such as blood pressure, lipid profile, and type 2 diabetes risk, therefore suggesting that tight control of the factors that are typically seen in clinical practice, may account for the beneficial effect of GFAT on vascular health. Our results are in line with prior epidemiologic and mendelian randomization evidence suggesting that common vascular factors are the predominant contributors that lie in the pathway between obesity and cardiovascular disease risk. In a study by Bakhtiyari et al. including 6280 individuals followed over a median of 13.9 years, the percentage of cardiovascular disease risk mediated by three traditional vascular risk factors – blood pressure, total cholesterol, and glucose – ranged from 46% to 52% and 66% in individuals in the overweight, visceral adiposity, and general adiposity categories, respectively^43^. Similarly, leveraging genetic data, Gill et al. demonstrated that the effect of obesity, either expressed as BMI or WHR, on cardiovascular risk through three vascular outcomes is mediated by up to 95% by common vascular risk factors and smoking^44^. Consistent with our results, blood pressure and diabetes were predominant mediators across outcomes.

Third, apart from common vascular risk factors, we found that adipose tissue-specific factors, such as adiponectin or leptin may mediate the metabolically protective role of GFAT in cardiovascular disease risk. Adiponectin is a hormone secreted predominantly from adipose tissue and is involved in biologic processes that interact with the insulin pathway, ultimately exerting anti-diabetic, anti-inflammatory, and anti-atherogenic effects^45^. Decreased serum adiponectin levels have been associated with increased risk of diabetes, cardiovascular diseases, obesity, and metabolic syndrome^46^. Our results provide further support to the suspected cardioprotective role of adiponectin, and suggest that targeting the adiponectin pathway may represent a viable therapeutic strategy to reduce cardiovascular disease risk in contexts where fat is not preferentially distributed in the gluteofemoral area. As thiazolidinediones have been shown to increase adiponectin levels, they might represent a reasonable treatment approach for reducing obesity-related cerebrovascular risk beyond their beneficial glycemic effects^47^. Interestingly, a recent meta-analysis found that GLP-1 RAs may be associated with increased adiponectin levels^48^, suggesting that the beneficial effects of these agents on cardiovascular disease risk^8^ might be partially attributable to adiponectin. Similar to adiponectin, leptin is another common adipokine predominantly secreted by the adipose tissue that acts primarily in the hypothalamus, regulating appetite through various neuropeptides^49^. Leptin function is pleiotropic and is affected by overall body mass and nutritional status^49^. There is contradictory evidence in the observational literature regarding a possible positive association of leptin with cardiovascular disease risk, including hypertension, CAD, and carotid artery disease, largely owning to uncontrolled confounding^49^. Pharmacologic targeting has been studied in conditions that represent human models of cardiometabolic dysregulation, such as congenital leptin deficiency (CLD) as well as general lipodystrophy (GL). In both these conditions, the phenotype is a state of dyslipidemia, insulin resistance, ectopic fat deposition in the liver and steatosis, and in the case of CLD, morbid obesity. In these states, treatment with leptin resulted in improvement of the cardiometabolic profile through correction of dyslipidemia and insulin resistance, weight loss, improvement in liver function and steatosis. In light of the above, our finding of the protective role of GFAT on cardiovascular risk potentially partially mediated through leptin, may suggest that in less severe spectrums of these phenotypes, such as patients who do not have preferentially distributed fat in the glutefemoral area, leptin administration or sensitization may exert a similar cardiometabolic benefit.

Our study has limitations. First, our analyses are based on individuals of predominantly European ancestry, limiting the generalizability of our results to other ancestries that might have divergent genetic backgrounds. As more GWAS of disease and mediator phenotypes from diverse populations become available, further analyses including individuals of non-European ancestries will be feasible and informative. Second, for purposes of power, males and females were grouped together in this set of analyses. However, it is important to note that a considerable amount of sex-dimorphism has been observed for adiposity metrics^7^, suggesting that the biological underpinnings of adiposity may, at least partly, be distinct between sexes. As more sex-stratified GWAS data of relevant traits become available, it would be interesting to evaluate whether the observed associations are driven by sex-specific factors. Third, although our results suggest a causal protective association between BMI-independent GFAT and cerebrovascular risk, the effect size estimates should be interpreted with caution as they reflect lifetime decreases, and not short-term adjustments^50^. Fourth, although we found that adipose-tissue specific factors may mediate the association between GFAT and stroke, our results do not directly provide evidence of targeting these factors to achieve a potential cardiometabolic benefit. Further analyses, including utilizing cis-instrumentation methodologies of specific molecular mediators, may provide further biological insight and replicative evidence.

In conclusion, our results suggest that independent of BMI, genetically proxied higher GFAT volume is associated with reduced cerebrovascular disease risk. Although common vascular risk factors predominantly mediate the protective association between GFAT and vascular risk, residual mediatory effect may be exerted by adipose-tissue specific factors, such as common adipokines. Therefore, targeting adipose-tissue specific pathways in the context of altered fat distribution profile states independent of BMI may provide additional cardiovascular benefit, beyond addressing traditional risk factors.

## Data Availability

All data produced in the present study are available upon reasonable request to the authors.

## Acknowledgements

E.P.M. has received research support from the American Academy of Neurology (AWARD NO A24-0242-001). M.K.G. has received research support from the German Research Foundation (DFG) with an Emmy Noether grant (GZ: GE 3461/2-1, ID 512461526), the framework of the Munich Cluster for Systems Neurology (EXC 2145 SyNergy, ID 390857198), the Fritz-Thyssen Foundation with a research grant (Ref. 10.22.2.024MN), and the Hertie Foundation with a research fellowship (Hertie Network of Excellence in Clinical Neuroscience, ID P1230035). J.R. is supported by NIH 1U24NS127780, R01NS093870, 1U01NS102289, and AHA 23BFHSCP1176239, 812095, 814722. C.D.A. is supported by NIH R01NS103924, U01NS069673, AHA 18SFRN34250007, AHA-Bugher 21SFRN812095, and the MGH McCance

Center for Brain Health.

## Author contributions

All authors contributed to the conception and design of the study. E.P.M. contributed to the acquisition and analysis of data, and drafting a significant portion of the manuscript or figures. M.K.G., E.M, and C.D.A. contributed to drafting a significant portion of the manuscript or figures.

