## Supplementary Material for "The role of the gluteofemoral adipose tissue in cerebrovascular disease risk: evidence from a mendelian randomization and mediation analysis"

**Supplementary Methods**

*Derivation of the local adiposity profiles in UK Biobank and baseline characteristics*

Briefly, from 40,032 participants in the UK Biobank with available whole body MRI data, 9040, 9041, and 7754 had available VAT, ASAT, and total adipose tissue (TAT - between the top of vertebrae T9 and the bottom of the thigh muscles) volumes, respectively. GFAT was calculated as the difference between TAT and the sum of VAT and ASAT. To reduce computational burden, the three-dimensional MRI images were transformed into two-dimensional projections of coronal and sagittal views. After verifying that the machine learning model had near-perfect estimation of adipose tissue volumes in a 20% held-out dataset of those with pre-calculated fat volumes (r^2^=0.991, 0.991, and 0.978 for VAT, ASAT, and GFAT, respectively), the local fat adiposity profiles were then calculated in the rest of the imaging dataset. The validity of MRI-derived GFAT distribution was further established by comparing it with DEXA-derived gynoid fat in 33,989 UK Biobank participants with available DEXA data, in which the correlation was very good (Pearson r = 0.96). Mean age of the whole cohort was 64.5 years, 51% were female, and 87% were of British ancestry. BMI was 26 kg/m^2^ (4.6) in females and 27.1 kg/m^2^ (3.8) in males. Males had higher VAT volumes (5 L (2.3) vs 2.6 L (1.5) in females), while females had higher ASAT (7.9 L (3.3) vs 5.9 L (2.5) in males) and GFAT (11.3 L (3.2) vs 9.3 (2.6) in males) volumes. Of note, given BMI and height had to be available, 37,641 subjects were available for the adjusted GWAS analysis. BMI- and height-adjusted local adiposity profiles were independent of BMI in both observational (Pearson r = 0 for VAT, ASAT, and GFAT) and genetic (r_g_ = -0.16, -0.07, and -0.04 for VAT, ASAT, and GFAT, respectively) correlation analyses, and had distinct genetic architecture between one another (r_g_ ranging from -0.18 to 0.35). Before each GWAS analysis, inverse-normal transformation was performed for each trait.

*Methodology used to calculate the phenotypic variance explained (R^2^) by each exposure instrument*

First, we calculated the phenotypic variance explained (R^2^) by each SNP included in the respective genetic instrument using the formula $\frac{2\beta^{2}\times MAF\times\left( 1-MAF \right)}{2\beta^{2}\times MAF\times\left( 1-MAF \right)+{se\left( \beta\right)}^{2}\times2N\times MAF\times\left( 1-MAF \right)}$, where *β* is the effect size for a given SNP, *MAF* is the minor allele frequency, *se(β)* is the standard error of the effect size, and *N* is the sample size. Because effective allele frequency (EAF), but not MAF was available in the exposure datasets, we considered EAF=MAF, if EAF was < 50%, and calculated the MAF as 1-EAF otherwise. As sample size, we considered the one depicted in **Table 1** for each respective phenotype. Finally, we calculated the total variance explained by each genetic proxy by summing the individual R^2^ of each SNP in the respective instrument.

**Supplementary Tables**

| **Supplementary Table 1.** GFAT SNPs | | | | | | | |
| --- | --- | --- | --- | --- | --- | --- | --- |
| **rsID** | **EA** | **OA** | **Beta** | **SE** | **P-value** | **F-statistic** | **R^2^** |
| rs72641832 | C | A | 0.0583338 | 0.00829183 | 5.2E-12 | 49.4925416802881 | 0.00131313066884325 |
| rs11205303 | T | C | -0.0392991 | 0.00724463 | 1.7E-08 | 29.4261056069521 | 0.00078114607794607 |
| rs2820446 | C | G | -0.0687621 | 0.00779316 | 7.1E-19 | 77.8522337079571 | 0.00206401385772771 |
| rs7588285 | C | G | 0.0526835 | 0.00931031 | 1.4E-08 | 32.0199725586093 | 0.000849944405357813 |
| rs13389219 | C | T | -0.0731577 | 0.00725328 | 3E-23 | 101.730505056115 | 0.00269536686124199 |
| rs78058190 | G | A | 0.115298 | 0.018548 | 3.7E-10 | 38.6410904260947 | 0.00102551641437776 |
| rs2943634 | A | C | 0.075414 | 0.00754268 | 4.8E-23 | 99.9660626888218 | 0.00264874149015622 |
| rs71304101 | G | A | -0.0618006 | 0.0108921 | 1.7E-09 | 32.1930514140745 | 0.000854534718364313 |
| rs2300669 | C | A | -0.0421979 | 0.00727957 | 4.4E-09 | 33.6023943377043 | 0.000891911054189508 |
| rs11920853 | G | A | -0.0371065 | 0.00726928 | 4.2E-08 | 26.0565666165314 | 0.000691760094671818 |
| rs62271373 | T | A | 0.122988 | 0.0154042 | 4.8E-15 | 63.7451549430428 | 0.00169064012184912 |
| rs13099700 | A | G | 0.0471756 | 0.00794479 | 7.9E-09 | 35.2590023936617 | 0.000935841384661401 |
| rs4450871 | A | G | -0.0383337 | 0.0071357 | 3.1E-08 | 28.8594674893519 | 0.000766115613313046 |
| rs874040 | G | C | 0.0448512 | 0.00778902 | 3E-08 | 33.1575548192275 | 0.000880114034958323 |
| rs13142096 | A | G | -0.0470762 | 0.00801245 | 8.4E-09 | 34.5201070377374 | 0.000916247657355871 |
| rs3822072 | G | A | 0.0478524 | 0.00712951 | 4.9E-12 | 45.049303801944 | 0.00119538409130615 |
| rs141171253 | C | A | 0.096977 | 0.0184705 | 2.9E-08 | 27.56640839766 | 0.000731814640854356 |
| rs6822892 | A | G | -0.0538768 | 0.00751089 | 8E-13 | 51.4541943738324 | 0.00136510597342618 |
| rs142369482 | G | GT | -0.0437893 | 0.00775141 | 9.1E-09 | 31.9135111135461 | 0.000847120865874405 |
| rs11429307 | G | GT | 0.0820665 | 0.0090607 | 3.1E-20 | 82.0367263157977 | 0.00217471162014294 |
| rs10044492 | C | T | -0.0478462 | 0.00805882 | 5.3E-09 | 35.2494213028675 | 0.000935587322100505 |
| rs1294437 | C | T | -0.0400038 | 0.00753026 | 4.1E-08 | 28.2216597585094 | 0.000749196784935383 |
| rs77318243 | T | A | -0.0678221 | 0.0121226 | 1.1E-08 | 31.3004740530754 | 0.000830861764723757 |
| rs199679345 | C | CA | 0.150135 | 0.0168141 | 1.6E-19 | 79.7290601421931 | 0.00211366699766255 |
| rs998584 | C | A | 0.07952 | 0.00711591 | 6.1E-31 | 124.879700806842 | 0.00330668057506348 |
| rs1317983 | T | C | 0.0579741 | 0.00770131 | 4.9E-14 | 56.6681219576566 | 0.0015032261882705 |
| rs72959041 | G | A | 0.195251 | 0.0168711 | 3.2E-32 | 133.936743199414 | 0.0035456510254389 |
| rs2982521 | A | T | -0.0551001 | 0.00735376 | 2.1E-14 | 56.1417402015108 | 0.00148928373902787 |
| rs11390479 | A | AG | 0.0531466 | 0.00818609 | 3.6E-11 | 42.1500352568814 | 0.00111853799954211 |
| rs1962883 | C | T | 0.0546516 | 0.007273 | 8.2E-14 | 56.4649030668912 | 0.00149784350783486 |
| rs111874795 | T | C | -0.102956 | 0.0170845 | 1E-09 | 36.3160642821433 | 0.000963870786873025 |
| rs1907218 | T | C | -0.0488068 | 0.00764797 | 3.6E-10 | 40.7256774271223 | 0.00108078058249648 |
| rs10501153 | C | T | -0.0439302 | 0.0076186 | 5.9E-09 | 33.2488031106067 | 0.000882533936757925 |
| rs71468663 | A | AC | 0.126663 | 0.0169041 | 1.1E-13 | 56.1455620363646 | 0.00148938497064635 |
| rs12814794 | G | A | -0.0721238 | 0.00826435 | 1.6E-18 | 76.1622556449964 | 0.00201929973227499 |
| rs4759309 | G | A | -0.0443202 | 0.00853853 | 4.2E-08 | 26.942457456917 | 0.000715262254829726 |
| rs150792771 | G | A | -0.15709 | 0.0278533 | 1.8E-08 | 31.8085422037944 | 0.000844336895354116 |
| rs7133378 | G | A | -0.0876372 | 0.00761231 | 5.6E-29 | 132.539111515686 | 0.00350878193129857 |
| rs60209099 | T | A | -0.0403624 | 0.00733363 | 3.1E-08 | 30.2911648302306 | 0.000804091494971384 |
| rs2955617 | C | A | -0.0418669 | 0.00747226 | 1.2E-08 | 31.3933498857983 | 0.000833325071604283 |
| rs8075019 | G | A | 0.0634833 | 0.0107754 | 2.3E-10 | 34.7098060394453 | 0.000921278091856448 |
| rs3786920 | T | C | -0.0509957 | 0.00731199 | 5E-12 | 48.6402655828339 | 0.00129054735572127 |
| rs1883711 | G | C | 0.127246 | 0.0208289 | 6.8E-10 | 37.3212007778678 | 0.000990521859479697 |
| rs2267373 | C | T | 0.0461893 | 0.00722912 | 1.4E-10 | 40.8236557706022 | 0.00108337792097147 |

*SNPs single nucleotide polymorphisms; GFAT gluteofemoral adipose tissue; EA effective allele; OA other allele; SE standard error; R^2^ variance explained*

| **Supplementary Table 2.** ASAT SNPs | | | | | | | |
| --- | --- | --- | --- | --- | --- | --- | --- |
| **rsID** | **EA** | **OA** | **Beta** | **SE** | **P-value** | **F-statistic** | **R^2^** |
| rs1779445 | T | C | -0.0493135 | 0.00906223 | 1.9E-08 | 29.6115740824441 | 0.000786065658217692 |
| rs3850625 | G | A | -0.0786112 | 0.0110023 | 1.8E-12 | 51.050721726135 | 0.00135441613678793 |
| rs6685593 | T | A | -0.0570031 | 0.00718973 | 5.2E-15 | 62.8596250743837 | 0.00166719337753369 |
| rs7538503 | A | G | -0.0474118 | 0.00787047 | 8.4E-10 | 36.2887118114014 | 0.000963145519545446 |
| rs2943647 | T | C | 0.0432106 | 0.00746638 | 5.8E-09 | 33.4934908794739 | 0.000889022990782404 |
| rs11709077 | G | A | -0.0696508 | 0.0110061 | 1.7E-10 | 40.0484128383112 | 0.00106282639483742 |
| rs7649153 | T | A | 0.0421552 | 0.00755389 | 2.7E-08 | 31.1430394243702 | 0.000826686164144649 |
| rs13322435 | A | G | 0.0570714 | 0.00731172 | 2.4E-15 | 60.9253382038336 | 0.00161597418851438 |
| rs55744247 | G | A | -0.0534533 | 0.0088675 | 5.1E-10 | 36.336797595716 | 0.000964420542537781 |
| rs3936510 | G | T | -0.0629779 | 0.00887846 | 5E-13 | 50.315415700678 | 0.0013349339269735 |
| rs1159619 | C | A | 0.0457682 | 0.0071716 | 1.2E-10 | 40.7282310736463 | 0.00108084827808031 |
| rs200015011 | C | CTCG | -0.0478625 | 0.00725275 | 7.3E-11 | 43.5497176107592 | 0.00115563852924074 |
| rs73221948 | G | T | -0.049661 | 0.00823067 | 2.9E-09 | 36.4049586462837 | 0.000966227867504167 |
| rs6474550 | G | T | 0.0450769 | 0.00757871 | 1.3E-09 | 35.3767151428149 | 0.000938962772622356 |
| rs17205757 | A | G | -0.0415864 | 0.00765851 | 3.2E-08 | 29.4858794417311 | 0.00078273159353707 |
| rs199797372 | TG | T | -0.060143 | 0.00721774 | 4.6E-17 | 69.4332841201696 | 0.0018412221253742 |
| rs8077609 | A | C | 0.0420664 | 0.00765749 | 1.1E-08 | 30.1785116671529 | 0.000801103465818207 |
| rs4444401 | A | G | -0.0396604 | 0.00725778 | 4.2E-08 | 29.8611535596497 | 0.000792685716366429 |
| rs2302209 | C | T | -0.0461843 | 0.00799643 | 3.4E-09 | 33.357727105021 | 0.000885422582294524 |

*SNPs single nucleotide polymorphisms; ASAT abdominal subcutaneous adipose tissue; EA effective allele; OA other allele; SE standard error; R^2^ variance explained*

| **Supplementary Table 3.** VAT SNPs | | | | | | | |
| --- | --- | --- | --- | --- | --- | --- | --- |
| **rsID** | **EA** | **OA** | **Beta** | **SE** | **P-value** | **F-statistic** | **R^2^** |
| rs12089366 | C | T | 0.0580131 | 0.00855551 | 9.4E-12 | 45.9790866271227 | 0.00122002579515422 |
| rs56006999 | C | T | 0.0536419 | 0.00922851 | 3.6E-09 | 33.786649778527 | 0.000896797375194311 |
| rs35932591 | C | T | 0.0606447 | 0.0107886 | 3.8E-08 | 31.5977036196136 | 0.000838745017484623 |
| rs3731861 | T | C | -0.0382281 | 0.00730702 | 4.7E-08 | 27.3706325746002 | 0.000726621091248656 |
| rs56082403 | T | C | -0.0556812 | 0.00723565 | 6.9E-14 | 59.2191364700762 | 0.00157079024542829 |
| rs30351 | G | A | 0.0705595 | 0.00808072 | 1.1E-16 | 76.2449128136192 | 0.00202148680238616 |
| rs72810972 | G | T | -0.0539032 | 0.00784907 | 2.3E-12 | 47.1620518896374 | 0.00125137574564406 |
| rs76072243 | T | C | -0.0550714 | 0.00738591 | 4.9E-14 | 55.5960851608035 | 0.00147483038084409 |
| rs80163833 | C | T | -0.0500918 | 0.0086022 | 5.8E-09 | 33.9089392760753 | 0.000900040377820932 |
| rs190594352 | C | T | -0.0532592 | 0.00891091 | 1.6E-09 | 35.7227729020853 | 0.000948139070306239 |
| rs185139895 | G | A | -0.100386 | 0.0176056 | 3.3E-09 | 32.5120712710889 | 0.000862995496931166 |
| rs998584 | C | A | -0.0570321 | 0.00708399 | 1.8E-15 | 64.8160927773618 | 0.00171899456088891 |
| rs72959041 | G | A | -0.117561 | 0.0167954 | 7.8E-13 | 48.994331945243 | 0.00129992940603115 |
| rs2982521 | A | T | 0.0596795 | 0.00732077 | 2.7E-16 | 66.4564023096398 | 0.00176242071596135 |
| rs1635851 | C | T | 0.040899 | 0.00720247 | 3.8E-08 | 32.2450060551904 | 0.000855912625790736 |
| rs11992444 | G | T | -0.0779981 | 0.00709912 | 1.3E-29 | 120.714381214768 | 0.00319673995719906 |
| rs4872393 | G | A | -0.0601652 | 0.00844375 | 2E-12 | 50.7715002625048 | 0.00134701814856729 |
| rs1329254 | C | T | 0.0418348 | 0.00733732 | 1.4E-08 | 32.5087651997015 | 0.000862907816798071 |
| rs11031796 | G | A | 0.0524669 | 0.00728182 | 5.1E-14 | 51.9147695739046 | 0.00137730843823759 |
| rs4307676 | G | A | 0.0535154 | 0.00949868 | 8.8E-09 | 31.7417624959174 | 0.000842565765349128 |
| rs7933253 | T | C | 0.0978848 | 0.0167983 | 1.3E-08 | 33.9546962342681 | 0.000901253803967069 |
| rs7133378 | G | A | 0.0458609 | 0.00758948 | 6.6E-10 | 36.5141424030831 | 0.000969122916790024 |
| rs57215491 | T | C | 0.0782434 | 0.0105443 | 1.3E-13 | 55.0630065124878 | 0.0014607097431627 |
| rs10406327 | C | G | -0.0705486 | 0.00709512 | 3.3E-24 | 98.8683552671302 | 0.002619732383177 |
| rs28451064 | G | A | -0.0687811 | 0.0106791 | 2.4E-11 | 41.4828826199023 | 0.00110085322002588 |

*SNPs single nucleotide polymorphisms; VAT visceral adipose tissue; EA effective allele; OA other allele; SE standard error; R^2^ variance explained*

| **Supplementary Table 4.** Univariable MR analyses between GFAT volume and ischemic stroke and stroke subtypes | | | | | | | | |
| --- | --- | --- | --- | --- | --- | --- | --- | --- |
| **Outcome** | **MR Method** | **OR** | **95% CI** | **FDR-p** | **N SNPs** | ***I^2^*** | ***Q*** | ***Q* p-value** |
| Ischemic stroke | IVW | 0.92 | 0.86-0.98 | 0.0084 | 38 | 45.5% | 67.8768 | 0.0015 |
| Ischemic stroke | Weighted Median | 0.91 | 0.85-0.98 | 0.0175 | 38 | - | - | - |
| Ischemic stroke | MR Egger | 0.95 | 0.80-1.14 | 0.6837 | 38 | - | - | - |
| Ischemic stroke | MR Egger intercept | 1.00 | 0.99-1.01 | 0.691 | 38 | - | - | - |
| Ischemic stroke | MR-APSS | 0.98 | 0.97-0.99 | <0.001 | 174 | - | - | - |
| Ischemic stroke | MR-APSS° | 0.97 | 0.96 0.99 | 0.001 | 34 | - | - | - |
| Large artery stroke | IVW | 0.80 | 0.66-0.96 | 0.019 | 34 | 38.6% | 53.7332 | 0.0127 |
| Large artery stroke | Weighted Median | 0.79 | 0.63-1.00 | 0.056 | 34 | - | - | - |
| Large artery stroke | MR Egger | 0.67 | 0.37-1.23 | 0.35 | 34 | - | - | - |
| Large artery stroke | MR Egger intercept | 1.01 | 0.98-1.05 | 0.691 | 34 | - | - | - |
| Large artery stroke | MR-APSS | 0.98 | 0.97-0.99 | 0. 0.0025 | 174 | - | - | - |
| Large artery stroke | MR-APSS° | 0.98 | 0.97-0.99 | 0.0117 | 34 | - | - | - |
| Cardioembolic stroke | IVW | 0.98 | 0.87-1.11 | 0.749 | 35 | 23.4% | 44.3665 | 0.1099 |
| Cardioembolic stroke | Weighted Median | 1.03 | 0.88-1.20 | 0.71 | 35 | - | - | - |
| Cardioembolic stroke | MR Egger | 1.05 | 0.72-1.53 | 0.786 | 35 | - | - | - |
| Cardioembolic stroke | MR Egger intercept | 1.00 | 0.97-1.02 | 0.691 | 35 | - | - | - |
| Cardioembolic stroke | MR-APSS | 1.00 | 0.99-1.01 | 0.81515 | 174 | - | - | - |
| Cardioembolic stroke | MR-APSS° | 0.99 | 0.98-1.01 | 0.4177 | 34 | - | - | - |
| Small vessel stroke | IVW | 0.77 | 0.67-0.88 | <0.001 | 34 | 0.0% | 28.7836 | 0.6772 |
| Small vessel stroke | Weighted Median | 0.80 | 0.66-0.97 | 0.0308 | 34 | - | - | - |
| Small vessel stroke | MR Egger | 0.68 | 0.44-1.04 | 0.1703 | 34 | - | - | - |
| Small vessel stroke | MR Egger intercept | 1.01 | 0.98-1.03 | 0.691 | 34 | - | - | - |
| Small vessel stroke | MR-APSS | 0.98 | 0.97-0.99 | 0.0025 | 174 | - | - | - |
| Small vessel stroke | MR-APSS° | 0.99 | 0.97-1.00 | 0.0285 | 34 | - | - | - |

*MR mendelian randomization; GFAT gluteofemoral adipose tissue; IVW inverse-variance weighted; OR odds ratio; CI confidence interval; SNPs single nucleotide polymorphisms; Q Cochran’s Q statistic; Q p-value Cochran’s Q p-value; MR-APSS° MR-APSS with p-value threshold < 5x10^-8^*

| **Supplementary Table 5.** Reverse IVW MR analyses between ischemic stroke and stroke subtypes and GFAT volume | | | | | | | | |
| --- | --- | --- | --- | --- | --- | --- | --- | --- |
| **Exposure** | **Outcome** | **Beta** | **95% CI** | **P-value** | **N SNPs** | ***I^2^*** | ***Q*** | ***Q* p-value** |
| Ischemic stroke | GFAT | -0.007 | (-0.098) – (0.083) | 0.874 | 26 | 57.6% | 58.9881 | 0.0001 |
| Large artery stroke | GFAT | 0.067 | (-0.001) – (0.134) | 0.053 | 3 | 74.8% | 7.9245 | 0.019 |
| Cardioembolic stroke | GFAT | 0.020 | (-0.019) – (0.060) | 0.306 | 9 | 5.4% | 8.4568 | 0.3902 |
| Small vessel stroke | GFAT |  |  |  |  |  |  |  |

*MR mendelian randomization; GFAT gluteofemoral adipose tissue; IVW inverse-variance weighted; CI confidence interval; SNPs single nucleotide polymorphisms*

| **Supplementary Table 6.** Univariable MR analyses between ASAT and VAT volumes on ischemic stroke and stroke subtypes | | | | | | |
| --- | --- | --- | --- | --- | --- | --- |
| **Exposure** | **Outcome** | **MR Method** | **OR** | **95% CI** | **P-value** | **N SNPs** |
| ASAT | Ischemic stroke | IVW | 1.04 | 0.92 – 1.16 | 0.548 | 17 |
| ASAT | Large artery stroke | IVW | 0.98 | 0.70 – 1.38 | 0.912 | 17 |
| ASAT | Cardioembolic stroke | IVW | 1.02 | 0.86 – 1.21 | 0.826 | 17 |
| ASAT | Small vessel stroke | IVW | 0.99 | 0.74 – 1.33 | 0.970 | 17 |
| VAT | Ischemic stroke | IVW | 0.96 | 0.87 – 1.04 | 0.31 | 20 |
| VAT | Large artery stroke | IVW | 1.16 | 0.84 – 1.59 | 0.368 | 19 |
| VAT | Cardioembolic stroke | IVW | 1.10 | 0.94 – 1.30 | 0.235 | 20 |
| VAT | Small vessel stroke | IVW | 1.18 | 0.98 – 1.44 | 0.088 | 19 |

*MR mendelian randomization; ASAT abdominal subcutaneous adipose tissue; VAT visceral adipose tissue; IVW inverse-variance weighted; CI confidence interval; SNPs single nucleotide polymorphisms*

| **Supplementary Table 7.** Multivariable MR between GFAT, ASAT, and VAT volumes and ischemic stroke and stroke subtypes | | | | | | | | |
| --- | --- | --- | --- | --- | --- | --- | --- | --- |
| **Exposure** | **Outcome** | **MR Method** | **OR** | **95% CI** | **P-value** | **N SNPs** | ***Q*** | ***Q* p-value** |
| GFAT | Ischemic stroke | IVW | 0.87 | 0.81 – 0.95 | 0.001 | 61 | 108.9711 | 0.0001 |
| ASAT | Ischemic stroke | IVW | 1.02 | 0.92 – 1.15 | 0.673 |  |  |  |
| VAT | Ischemic stroke | IVW | 0.90 | 0.82 – 0.99 | 0.034 |  |  |  |
| GFAT | Ischemic stroke | Weighted median | 0.90 | 0.82 – 0.99 | 0.027 | - | - | - |
| ASAT | Ischemic stroke | Weighted median | 1.03 | 0.90 – 1.16 | 0.699 |  |  |  |
| VAT | Ischemic stroke | Weighted median | 0.99 | 0.89 – 1.10 | 0.865 |  |  |  |
| GFAT | Ischemic stroke | MR Egger | 0.88 | 0.79 – 0.99 | 0.026 | 61 | 108.8724 | < 0.001 |
| ASAT | Ischemic stroke | MR Egger | 1.03 | 0.91 – 1.16 | 0.640 |  |  |  |
| VAT | Ischemic stroke | MR Egger | 0.90 | 0.82 – 0.99 | 0.035 |  |  |  |
| GFAT | Large artery stroke | IVW | 0.71 | 0.55 – 0.92 | 0.01 | 56 | 99.0239 | 0.0001 |
| ASAT | Large artery stroke | IVW | 1.07 | 0.75 – 1.51 | 0.725 |  |  |  |
| VAT | Large artery stroke | IVW | 0.77 | 0.57 – 1.03 | 0.081 |  |  |  |
| GFAT | Large artery stroke | Weighted median | 0.80 | 0.60 – 1.06 | 0.122 | - | - | - |
| ASAT | Large artery stroke | Weighted median | 1.00 | 0.67 – 1.51 | 0.981 |  |  |  |
| VAT | Large artery stroke | Weighted median | 0.88 | 0.63 – 1.24 | 0.471 |  |  |  |
| GFAT | Large artery stroke | MR Egger | 0.79 | 0.49 – 1.01 | 0.056 | 56 | 98.9872 | 0.0001 |
| ASAT | Large artery stroke | MR Egger | 1.06 | 0.73 – 1.54 | 0.773 |  |  |  |
| VAT | Large artery stroke | MR Egger | 0.77 | 0.56 – 1.05 | 0.098 |  |  |  |
| GFAT | Small vessel stroke | IVW | 0.68 | 0.57 – 0.80 | < 0.001 | 57 | 54.0067 | 0.4741 |
| ASAT | Small vessel stroke | IVW | 1.07 | 0.85 – 1.36 | 0.559 |  |  |  |
| VAT | Small vessel stroke | IVW | 0.84 | 0.69 – 1.03 | 0.095 |  |  |  |
| GFAT | Small vessel stroke | Weighted median | 0.74 | 0.58 – 0.95 | 0.019 | - | - | - |
| ASAT | Small vessel stroke | Weighted median | 1.16 | 0.82 – 1.64 | 0.392 |  |  |  |
| VAT | Small vessel stroke | Weighted median | 0.92 | 0.70 – 1.22 | 0.570 |  |  |  |
| GFAT | Small vessel stroke | MR Egger | 0.75 | 0.59 – 0.96 | 0.023 | 57 | 52.4761 | 0.4945 |
| ASAT | Small vessel stroke | MR Egger | 1.13 | 0.88 – 1.45 | 0.342 |  |  |  |
| VAT | Small vessel stroke | MR Egger | 0.82 | 0.67 – 1.01 | 0.058 |  |  |  |

*MR mendelian randomization; GFAT gluteofemoral adipose tissue; ASAT abdominal subcutaneous adipose tissue; VAT visceral adipose tissue; IVW inverse-variance weighted; OR odds ratio; CI confidence interval; SNPs single nucleotide polymorphisms; Q Cochran’s Q statistic; Q p-value Cochran’s Q p-value*

| **Supplementary Table 8.** Univariable MR analyses between GFAT volume and positive controls | | | | | | | | |
| --- | --- | --- | --- | --- | --- | --- | --- | --- |
| **Outcome** | **MR Method** | **Beta** | **95% CI** | **FDR-p** | **N SNPs** | ***I^2^*** | ***Q*** | ***Q* p-value** |
| Coronary artery disease | IVW | -0.198 | (-0.272) – (-0.123) | <0.001 | 44 | 83.8% | 266.1746 | < 0.001 |
| Coronary artery disease | Weighted Median | -0.208 | (-0.266) – (-0.150) | <0.001 | 44 | - | - | - |
| Coronary artery disease | MR Egger | -0.429 | (-0.630) – (-0.228) | <0.001 | 44 | - | - | - |
| Coronary artery disease | MR Egger intercept | 0.015 | 0.003 – 0.028 | 0.112 | 44 | - | - | - |
| Coronary artery disease | MR-APSS | -0.047 | (-0.072) – (-0.022) | <0.001 | 174 | - | - | - |
| Coronary artery disease | MR-APSS° | -0.0606 | (-0.103) – (-0.018) | 0.0117 | 34 | - | - | - |
| MRI-confirmed lacunar stroke | IVW | -0.245 | (-0.405) – (-0.086) | 0.0053 | 32 | 15.8% | 36.8274 | 0.2172 |
| MRI-confirmed lacunar stroke | Weighted Median | -0.288 | (-0.503) – (-0.072) | 0.0175 | 32 | - | - | - |
| MRI-confirmed lacunar stroke | MR Egger | -0.136 | (-0.625) – (0.353) | 0.6837 | 32 | - | - | - |
| MRI-confirmed lacunar stroke | MR Egger intercept | -0.007 | (-0.036) – (0.022) | 0.691 | 32 | - | - | - |
| MRI-confirmed lacunar stroke | MR-APSS | -0.0291 | (-0.053) – (-0.006) | 0.022 | 172 | - | - | - |
| MRI-confirmed lacunar stroke | MR-APSS° | -0.0306 | (-0.057) – (-0.005) | 0.0297 | 34 | - | - | - |
| Mean carotid IMT | IVW | -0.073 | (-0.272) – (-0.123) | 0.0023 | 44 | 31.8% | 63.0857 | 0.0245 |
| Mean carotid IMT | Weighted Median | -0.101 | (-0.152) – (-0.050) | <0.001 | 44 | - | - | - |
| Mean carotid IMT | MR Egger | -0.118 | (-0.234) – (-0.003) | 0.1575 | 44 | - | - | - |
| Mean carotid IMT | MR Egger intercept | 0.003 | (-0.004) – (0.010) | 0.691 | 44 | - | - | - |
| Mean carotid IMT | MR-APSS | -0.0808 | (-0.151) – (-0.011) | 0.0278 | 174 | - | - | - |
| Mean carotid IMT | MR-APSS° | -0.0284 | (-0.102) – (0.045) | 0.451 | 34 | - | - | - |

*MR mendelian randomization; GFAT gluteofemoral adipose tissue; CAD coronary artery disease; IVW inverse-variance weighted; CI confidence interval; SNPs single nucleotide polymorphisms; IMT intima media thickness; MR-APSS° MR-APSS with p-value threshold < 5x10^-8^*

| **Supplementary Table 9.** Univariable MR between GFAT volume and mediators | | | | |
| --- | --- | --- | --- | --- |
| **Exposure** | **Mediator** | **Beta (95% CI)** | **P-value** | **N SNPs** |
| **GFAT** | **Systolic blood pressure** | -1.866 [(-2.411) – (-1.320)] | <0.001 | 31 |
| **GFAT** | **Type 2 diabetes** | -0.372 [(-0.509) – (-0.235)] | <0.001 | 34 |
| **GFAT** | **LDL** | -0.091 [(-0.134) – (-0.048)] | <0.001 | 44 |
| **GFAT** | **Fasting insulin** | -0.031 [(-0.076) - (0.014)] | 0.176 | 44 |
| **GFAT** | **CRP** | 0.049 (0.011 – 0.087) | 0.011 | 42 |
| **GFAT** | **Adiponectin** | 0.417 (0.268 – 0.566) | <0.001 | 38 |
| **GFAT** | **Leptin** | 0.293 (0.100 – 0.486) | 0.003 | 6 |

*MR mendelian randomization; GFAT glutefemoral adipose tissue; SBP systolic blood pressure; LDL low density lipoprotein; CRP c-reactive protein; CI confidence interval; SNPs single nucleotide polymorphisms*

| **Supplementary Table 10.** Univariable MR between mediators and outcomes | | | | |
| --- | --- | --- | --- | --- |
| **Outcome** | **Mediator** | **Beta (95% CI)** | **P-value** | **N SNPs** |
| **Ischemic stroke** | **Systolic blood pressure** | 0.027 (0.024 – 0.030) | <0.001 | 783 |
| **Ischemic stroke** | **Type 2 diabetes** | 0.096 (0.074 – 0.118) | <0.001 | 241 |
| **Ischemic stroke** | **LDL** | 0.117 (0.079 0.155) | <0.001 | 766 |
| **Ischemic stroke** | **Fasting insulin** |  |  |  |
| **Ischemic stroke** | **CRP** | 0.036 [(-0.008) – (0.080)] | 0.110 | 393 |
| **Ischemic stroke** | **Adiponectin** | -0.032 [(-0.059) – (-0.005)] | 0.019 | 36 |
| **Ischemic stroke** | **Leptin** | 0.053 [(-0.198) – (0.304)] | 0.681 | 4 |
| **Large artery stroke** | **Systolic blood pressure** | 0.046 (0.039 – 0.053) | <0.001 | 740 |
| **Large artery stroke** | **Type 2 diabetes** | 0.175 (0.109 – 0.242) | <0.001 | 226 |
| **Large artery stroke** | **LDL** | 0.337 (0.216 – 0.458) | <0.001 | 602 |
| **Large artery stroke** | **Fasting insulin** |  |  |  |
| **Large artery stroke** | **CRP** |  |  |  |
| **Large artery stroke** | **Adiponectin** | -0.052 [(-0.128) – (0.023)] | 0.176 | 30 |
| **Large artery stroke** | **Leptin** |  |  |  |
| **Small vessel stroke** | **Systolic blood pressure** | 0.037 (0.030 – 0.043) | <0.001 | 736 |
| **Small vessel stroke** | **Type 2 diabetes** | 0.155 (0.102 – 0.208) | <0.001 | 225 |
| **Small vessel stroke** | **LDL** | 0.056 [(-0.055) – (0.167)] | 0.322 | 608 |
| **Small vessel stroke** | **FI** |  |  |  |
| **Small vessel stroke** | **CRP** |  |  |  |
| **Small vessel stroke** | **Adiponectin** | -0.028 [(-0.113) – (0.058)] | 0.525 | 28 |
| **Small vessel stroke** | **Leptin** |  |  |  |
| **Coronary artery disease** | **Systolic blood pressure** | 0.010 (0.007 – 0.014) | <0.001 | 785 |
| **Coronary artery disease** | **Type 2 diabetes** | 0.041 (0.010 – 0.072) | 0.009 | 244 |
| **Coronary artery disease** | **LDL** | 0.075 (0.042 – 0.107) | <0.001 | 860 |
| **Coronary artery disease** | **Fasting insulin** |  |  |  |
| **Coronary artery disease** | **CRP** | 0.006 [(-0.025) – (0.037)] | 0.708 | 443 |
| **Coronary artery disease** | **Adiponectin** | -0.027 [(-0.052) – (-0.001)] | 0.038 | 36 |
| **Coronary artery disease** | **Leptin** | -0.303 [(-0.454) – (-0.153)] | <0.001 | 4 |
| **MRI-confirmed lacunar stroke** | **Systolic blood pressure** | 0.035 (0.028 – 0.042) | <0.001 | 773 |
| **MRI-confirmed lacunar stroke** | **Type 2 diabetes** | 0.100 (0.045 – 0.154) | <0.001 | 236 |
| **MRI-confirmed lacunar stroke** | **LDL** | 0.044 [(-0.057) – (0.145)] | 0.393 | 704 |
| **MRI-confirmed lacunar stroke** | **Fasting insulin** |  |  |  |
| **MRI-confirmed lacunar stroke** | **CRP** | -0.027 [(-0.125) – (0.070)] | 0.586 | 373 |
| **MRI-confirmed lacunar stroke** | **Adiponectin** | 0.014 [(-0.056) – (0.084)] | 0.698 | 34 |
| **MRI-confirmed lacunar stroke** | **Leptin** | -0.315 [(-0.767) – (0.138)] | 0.173 | 4 |
| **Mean carotid IMT** | **Systolic blood pressure** | 0.018 (0.016 – 0.021) | <0.001 | 785 |
| **Mean carotid IMT** | **Type 2 diabetes** | 0.028 (0.008 – 0.047) | 0.005 | 244 |
| **Mean carotid IMT** | **LDL** | 0.114 (0.084 – 0.143) | <0.001 | 861 |
| **Mean carotid IMT** | **Fasting insulin** |  |  |  |
| **Mean carotid IMT** | **CRP** | -0.018 [(-0.049) – (0.013)] | 0.250 | 447 |
| **Mean carotid IMT** | **Adiponectin** | -0.012 [(-0.030) – (0.006)] | 0.190 | 36 |
| **Mean carotid IMT** | **Leptin** | 0.053 [(-0.067) – (0.172)] | 0.388 | 4 |

*MR mendelian randomization; LDL low density lipoprotein; CRP c-reactive protein; CI confidence interval; SNPs single nucleotide polymorphisms*

| **Supplementary Table 11.** Multivariable MR between mediators and outcomes, adjusting for GFAT | | | | |
| --- | --- | --- | --- | --- |
| **Outcome** | **Mediator** | **Beta (95% CI)** | **P-value** | **N SNPs** |
| **Ischemic stroke** | **Systolic blood pressure** | 0.027 (0.024 – 0.030) | <2e^-16^ | 783 |
| **Ischemic stroke** | **Type 2 diabetes** | 0.094 (0.070 – 0.118) | 1.5e^-13^ | 241 |
| **Ischemic stroke** | **LDL** | 0.116 (0.078 – 0.155) | 3.79e^-09^ | 766 |
| **Ischemic stroke** | **Fasting insulin** |  |  |  |
| **Ischemic stroke** | **CRP** |  |  |  |
| **Ischemic stroke** | **Adiponectin** | -0.031 [(-0.059) – (-0.003)] | 0.0323 | 36 |
| **Ischemic stroke** | **Leptin** |  |  |  |
| **Large artery stroke** | **Systolic blood pressure** | 0.046 (0.038 – 0.053) | <2e^-16^ | 740 |
| **Large artery stroke** | **Type 2 diabetes** | 0.172 (0.099 – 0.245) | 5.92e^-06^ | 226 |
| **Large artery stroke** | **LDL** | 0.330 (0.209 – 0.452) | 1.27e^-07^ | 602 |
| **Large artery stroke** | **Fasting insulin** |  |  |  |
| **Large artery stroke** | **CRP** |  |  |  |
| **Large artery stroke** | **Adiponectin** |  |  |  |
| **Large artery stroke** | **Leptin** |  |  |  |
| **Small vessel stroke** | **Systolic blood pressure** | 0.036 (0.029 – 0.043) | <2e^-16^ | 736 |
| **Small vessel stroke** | **Type 2 diabetes** | 0.153 (0.095 – 0.212) | 4.85e^-07^ | 225 |
| **Small vessel stroke** | **LDL** |  |  |  |
| **Small vessel stroke** | **Fasting insulin** |  |  |  |
| **Small vessel stroke** | **CRP** |  |  |  |
| **Small vessel stroke** | **Adiponectin** |  |  |  |
| **Small vessel stroke** | **Leptin** |  |  |  |
| **Coronary artery disease** | **Systolic blood pressure** | 0.010 (0.007 – 0.014) | 1.18e^-07^ | 785 |
| **Coronary artery disease** | **Type 2 diabetes** | 0.035 (0.002 – 0.069) | 0.0405 | 244 |
| **Coronary artery disease** | **LDL** | 0.072 (0.039 – 0.105) | 1.99e^-05^ | 856 |
| **Coronary artery disease** | **Fasting insulin** |  |  |  |
| **Coronary artery disease** | **CRP** |  |  |  |
| **Coronary artery disease** | **Adiponectin** | -0.022 [(-0.045) – (0.001)] | 0.06495 | 36 |
| **Coronary artery disease** | **Leptin** | -0.279 [(-0.715) – (0.156)] | 0.110 | 4 |
| **MRI-confirmed lacunar stroke** | **Systolic blood pressure** | 0.034 (0.028 – 0.041) | <2e^-16^ | 773 |
| **MRI-confirmed lacunar stroke** | **Type 2 diabetes** | 0.092 (0.032 – 0.151) | 0.00268 | 236 |
| **MRI-confirmed lacunar stroke** | **LDL** |  |  |  |
| **MRI-confirmed lacunar stroke** | **Fasting insulin** |  |  |  |
| **MRI-confirmed lacunar stroke** | **CRP** |  |  |  |
| **MRI-confirmed lacunar stroke** | **Adiponectin** |  |  |  |
| **MRI-confirmed lacunar stroke** | **Leptin** |  |  |  |
| **Mean carotid IMT** | **Systolic blood pressure** | 0.018 (0.015 – 0.021) | <2e-^16^ | 785 |
| **Mean carotid IMT** | **Type 2 diabetes** | 0.019 [(-0.002) – (0.040)] | 0.0733 | 244 |
| **Mean carotid IMT** | **LDL** | 0.111 (0.081 – 0.140) | 3.83e^-13^ | 859 |
| **Mean carotid IMT** | **Fasting insulin** |  |  |  |
| **Mean carotid IMT** | **CRP** |  |  |  |
| **Mean carotid IMT** | **Adiponectin** |  |  |  |
| **Mean carotid IMT** | **Leptin** |  |  |  |

*MR mendelian randomization; LDL low density lipoprotein; CRP c-reactive protein; CI confidence interval; SNPs single nucleotide polymorphisms*

| **Supplementary Table 12.** Mediation analysis between GFAT and cardiovascular outcomes | | |
| --- | --- | --- |
| **Outcome** | **Mediator** | **Percent mediated (95% CI)** |
| **Ischemic stroke** | **Systolic blood pressure** | 58.25924 (30.07796 - 170.23175) |
| **Ischemic stroke** | **Type 2 diabetes** | 40.208 (18.06195 - 141.61559) |
| **Ischemic stroke** | **LDL** | 12.18249 (4.43318 - 41.57389) |
| **Ischemic stroke** | **Adiponectin** | 14.86821 (1.81444 - 57.02112) |
| **Large artery stroke** | **Systolic blood pressure** | 37.12345 (18.23365 - 177.97104) |
| **Large artery stroke** | **Type 2 diabetes** | 27.88976 (11.48364 - 145.83877) |
| **Large artery stroke** | **LDL** | 13.13023 (4.248217 - 47.879815) |
| **Small vessel stroke** | **Systolic blood pressure** | 25.15407 (14.61573 - 53.10214) |
| **Small vessel stroke** | **Type 2 diabetes** | 21.45188 (10.73713 - 46.85328) |
| **Coronary artery disease** | **Systolic blood pressure** | 9.690949 (4.912001 - 17.235187) |
| **Coronary artery disease** | **Type 2 diabetes** | 6.660303 (1.423170 - 17.323812) |
| **Coronary artery disease** | **LDL** | 3.320397 (1.451556 - 6.944223) |
| **Coronary artery disease** | **Adiponectin** | 4.620697 (0.7857041 - 13.5269011) ***** |
| **MRI-confirmed lacunar stroke** | **Systolic blood pressure** | 26.24815 (14.22290 - 76.52831) |
| **MRI-confirmed lacunar stroke** | **Type 2 diabetes** | 13.93406 (5.588152 - 48.970090) |
| **Mean carotid IMT** | **Systolic blood pressure** | 45.83203 (24.81368 - 109.53577) |
| **Mean carotid IMT** | **Type 2 diabetes** | 9.636329 (4.108523 - 39.855442) ***** |
| **Mean carotid IMT** | **LDL** | 13.83325 (6.08596 - 34.05125) |

*Suggestive mediator.

95% CI were generated after bootstrapping over 1000 iterations.

*GFAT gluteofemoral adipose tissue; LDL low density lipoprotein; CI confidence interval; IMT intima media thickness*
